# The characteristics of oxygen concentration and the role of correction factor in real-time GI Breath Test

**DOI:** 10.1101/2020.10.22.20213876

**Authors:** Siu Man Lee, Imogen HE Falconer, Trudi Madden, Peter O Laidler

**Author notes:** Corresponding author: Dr Siu Man Lee.

## Abstract

A high quality end-expiratory breath sample is required for a reliable GI breath test result. Oxygen (O_2_) concentration in the breath sample can be used as a quality marker. This study investigated the characteristics of oxygen concentration in breath sample and the issues with using a correction factor in real-time breath test. The results indicated 95.4% of 564 patients were able to achieve an O_2_ concentration below 14% in their end-expiratory breath. A further 293 samples were studied and revealed that the distribution of O_2_ concentration was between 16.5% and 9.5%. Applying a correction factor to predict the end-expiratory H_2_ and CH_4_ values led to an average error of −36.4% and −12.8% respectively. The correction factor algorithm based on limiting O_2_ at 14% would have resulted in false negative result for 50% of the positive cases. This study has also indicated the continuous O_2_ measurement is essential to ensure breath sample quality by preventing secondary breathing during real-time breath collection.

## INTRODUCTION

Breath test (BT) has been widely used as a diagnostic tool to identify conditions related to the Gastrointestinal (GI) tract. It is a non-invasive, low cost and functional diagnostic test. Depending on the type of carbohydrate administered during the test, it can provide useful information to assist diagnosis of conditions like lactose mal-digestion, using lactose, and Small Intestine Bacterial Overgrowth (SIBO), using glucose or lactulose. The bacterial colonies in the digestive tract metabolise the carbohydrate and produce hydrogen or methane. These trace gases are absorbed in the intestine, returned to the lungs and equilibrated with air in the alveoli. The concentration of these trace gases can then be detected in the breath.

However, although GI breath test is simple and well tolerated by patients, there are a number of uncertainties within the test result, mainly related to the quality of the breath samples collected as well as the patient preparation procedures. Such uncertainties can adversely affect the accuracy of the result. There are criticisms among clinicians who do not consider Hydrogen Breath Test (HBT) as a reliable diagnostic test [1,2]. Significant effort has been spent on standardising and refining the protocol in order to make the test more reliable, such as the Rome Consensus and North American Consensus [3,4]. A common understanding in the test protocol is that alveolar air or end-expiratory breath is critical to the accuracy of the test result [5-10]. In addition, it is well-known that methane can be produced instead of hydrogen when the patient possess methanogenic bacteria that converts hydrogen (H_2_) to methane (CH_4_). It has been estimated between 5% – 15% of the population is affected [11].

Traditionally, only H_2_ concentration is measured. This may be due to the availability of a cost-effective detection system with precision down to part-per-million (ppm). More modern breath analysers are now commercially available and they are able to concurrently measure both H_2_and CH_4_. This equipment often includes measurement of oxygen (O_2_) or carbon dioxide (CO_2_) as a quality indicator of the breath sample. The rational that defined the CO_2_ concentration of an end-tidal breath as 5% was published in the seventies [12]. This value was widely adopted in subsequent research in breath tests. O_2_ concentration of alveolar air was approximated as 14% but it was estimated mathematically using the alveolar gas equation [13]. This equation has been widely used in studies on sustainable breathing, such as safety limit for hypoxemia [14]. However, the application of the alveolar gas equation can be limited by the conditions that the equation was derived. It may not be applicable to the single and maximum exhalation in GI breath test conditions.

The main technologies employed in the breath analysers are gas chromatography, electro-chemical sensing and opto-electronic sensing. There are a range of different types of analyser designs commercially available. However, they can be categorised into either Point-of-Care (POC) systems or laboratory systems. POC analysers can take either real-time measurement or can use a collection bag to collect breath sample from patients. Real-time measurement collects a breath sample and concurrently analyses the trace H_2_/CH_4_ concentration so no sample storage is required. It avoids volume normalisation, sample contamination and storage issues which can affect its accuracy. Laboratory based analysers require breath samples to be collected in a vessel and the batch of breath samples will be analysed at the same time.

This study demonstrated the characteristics of end-expiratory oxygen concentration in breath sample during real-time GI breath measurement. The results also illustrated the issues with the correction factor based on O_2_ concentration in the breath that may lead to an incorrect diagnostic result. The findings in this study may be used to guide future development of GI breath analysers and may help to reduce uncertainties in the GI breath test result. It may also help to refine the breath collection protocol and further improve the accuracy of the test result.

## METHODS

The clinical data in this study was collected via an audit of Combined H_2_/CH_4_ Breath Test (CBT) results at the Royal United Hospitals Bath NHS Foundation Trust, UK. The patients had followed the standard preparation protocol included a fasting period of 12 hours prior to the test as described in the North American Consensus [3].

Breath samples were collected in real-time measurement. In order to obtain an end-expiratory breath sample, every patient undergoing CBT test followed a strict instruction during each breath sample collection. Patients were asked to breathe in normally; hold their breath for 5 seconds and breathe out completely. When the patient was breathing into the analyser, the flow-rate was strictly monitored and maintained at optimum level as specified by the manufacturer. There was no noticeable leakage around the mouth piece and nose while the patients were encouraged to breathe out completely.

A breath sample was taken every 20 minutes, and the hydrogen, methane, and oxygen level,as well as the correction factor from the breath analyser were recorded.

CBT was performed by the GastroCH_4_ECK Gastrolyzer (Version 1) (software version: V11.0), manufactured by Bedfont Scientific Ltd. UK, see Figure 1. This instrument provides continuous real-time measurement of H_2_, CH_4_ and O_2_ concentration, in parts per million (ppm), during an episode of breath sample collection. It also provides a ratiometric correction factor according to the _EE_O_2_. Typically in a breath collection episode, the hydrogen and methane concentration will rise from zero; while the oxygen concentration will drop from the atmospheric concentration (20.9%) to _*EE*_*O*_*2*_ which is typically below 14%. The manufacturer regards the ideal _*EE*_*O*_*2*_ as 13.9% (< 14%) [15]. The _*EE*_*O*_*2*_ value is used to produce the ratiometric Correction Factor (CF). The CF is applied to the H_2_ and CH_4_ measurement and attempts to compensate for the non-expiry breath sample. The predicted hydrogen (_*CF*_*H*_*2*_) and predicted methane (_*CF*_*CH*_*4*_) values are expressed as the product of the actual H_2_/CH_4_measurement and the CF. When the _*EE*_*O*_*2*_ has not yet reached the manufacturer specified compliant level during a breath collection episode, a predicted value will be calculated. If _*EE*_*O*_*2*_is below 14%, CF will be equal to 1 so _CF_H_2_/_CF_CH_4_remain the same as the actual H_2_/CH_4_measured. The relationship between CF and the O_2_ concentration in the breath sample is shown in Figure 2.

**Figure 1:**
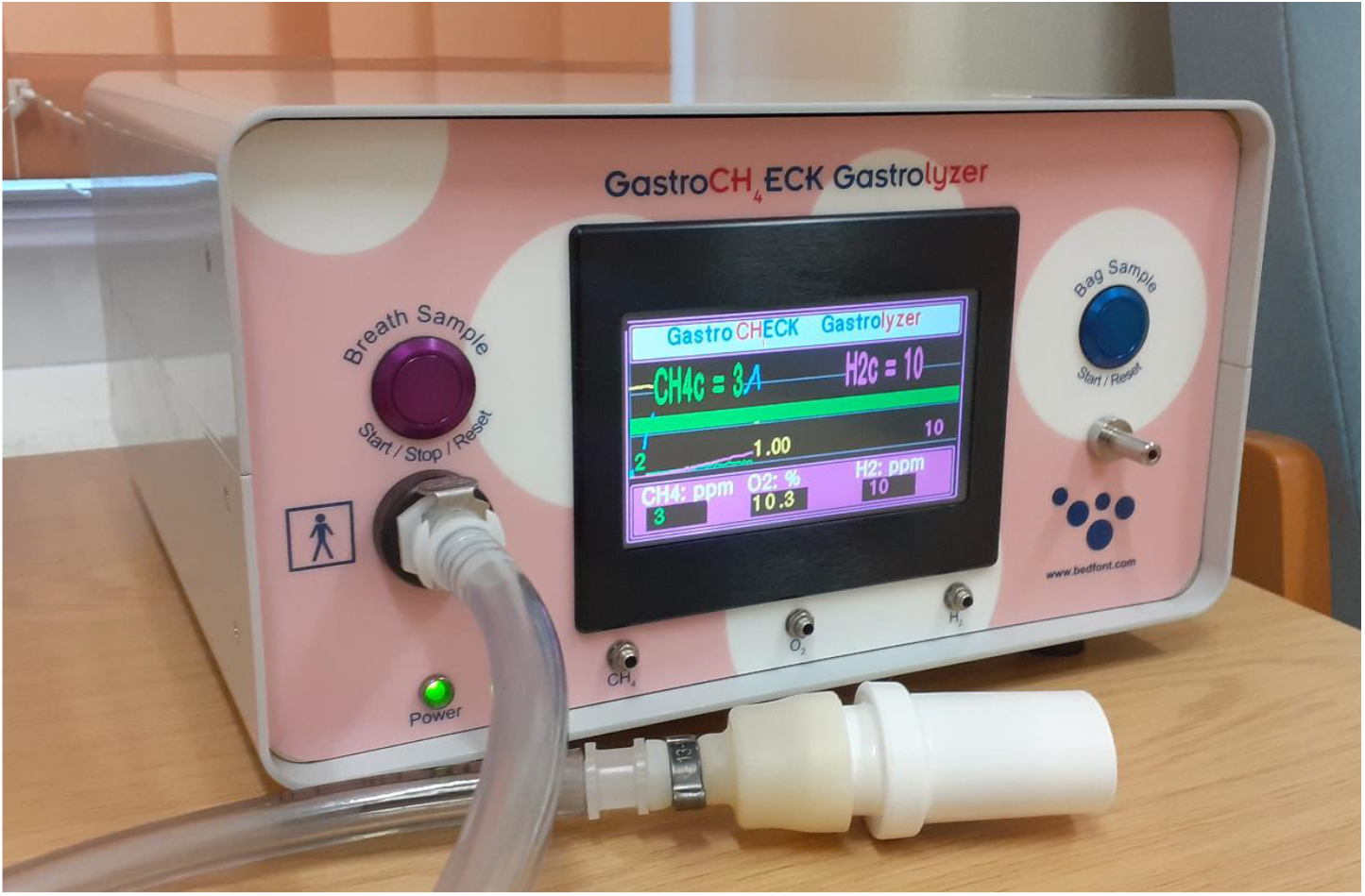
Real-time breath measurement setup on GastroCH_4_ECK Gastrolyzer (Ver. 1), software version: V11.0, manufactured by Bedfont Scientific Ltd. UK.

**Figure 2:**
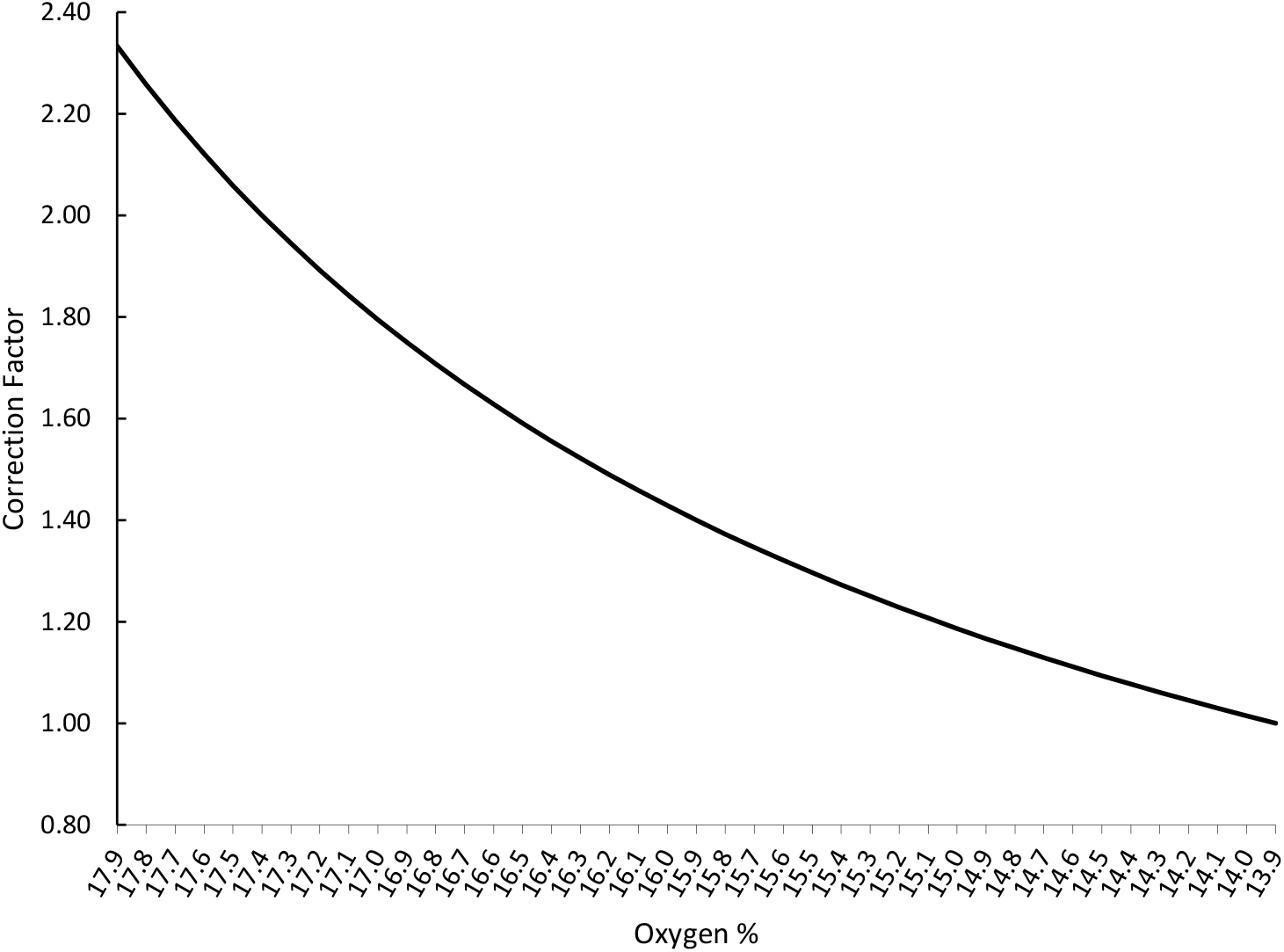
Correction Factor (CF), in GastroCH_4_ECK, in relation to the percentage of oxygen in breath sample. The CF value is 1 when oxygen level is below 13.9.

The characteristics of the breath samples collected from 564 patients were analysed. The analysis has taken into account for the demographics, _*EE*_*O*_*2*_ and the end-expiratory hydrogen (_*EE*_*H*_*2*_) and end-expiratory methane (_*EE*_*CH*_*4*_) concentrations. The data was analysed per patient. Typically, there were 5 samples collected for Glucose Breath Test and 6 samples collected for Lactose mal-digestion investigation.

A further 46 patients were studied in greater detail where extra hydrogen and methane readings were recorded when O_2_ was at 15%,as well as _*EE*_*H*_*2*_ and _*EE*_*CH*_*4*_values. This data was collected over a period of 12 weeks and it was analysed per breath sample.

In order to evaluate the efficacy of the CF, the diagnostic test result based on _*EE*_*H*_*2*_ and _*EE*_*CH*_*4*_was compared with the predicted result using _*CF*_*H*_*2*_and _*CF*_*CH*_*4*_.

## RESULTS

The results indicated that an average of 95.4% of the 564 patients who had undergone the Combined H_2_/CH_4_ Breath Test (CBT) achieved _*EE*_*O*_*2*_ below 14% (Table 1). There is no obvious difference between male and female, achieving 96.3% and 94.9% respectively. The result showed that none of the age groups between 20 −80 years old had any difficulty delivering _*EE*_*O*_*2*_ below 14% in their end-expiratory breath samples (Table 2). The 81 – 90 years old group has an obvious reduction in delivering _*EE*_*O*_*2*_ below 14% but still achieves a 73.3% success rate.

**Table 1:** The demographic of the patients who were able to deliver _*EE*_*O*_*2*_ below 14%

| | No. of Patients | No. of patients below 14% $\dot{V}O_2$ | % of patients below 14% $\dot{V}O_2$ | No. of patients above 14% $\dot{V}O_2$ | % of patients above 14% $\dot{V}O_2$ |
| --- | --- | --- | --- | --- | --- |
| Male | 191 | 184 | 96.3 | 7 | 3.7 |
| Female | 373 | 354 | 94.9 | 19 | 5.1 |
| Total | 564 | 538 | 95.4 | 26 | 4.6 |

**Table 2:** The distribution of patients who were able to deliver _*EE*_*O*_*2*_ below 14% by age group

| Age group | No. of patients | No. of patients below 14% $\dot{V}O_2$ | % of patients below 14% $\dot{V}O_2$ | Male | | Female | |
| --- | --- | --- | --- | --- | --- | --- | --- |
| | | | | No. of patients below 14% $\dot{V}O_2$ | % of patients below 14% $\dot{V}O_2$ | No. of patients below 14% $\dot{V}O_2$ | % of patients below 14% $\dot{V}O_2$ |
| 11-20 | 19 | 17 | 89.5 | 7 | 100 | 10 | 83.3 |
| 21-30 | 76 | 73 | 96.1 | 24 | 100 | 49 | 94.2 |
| 31-40 | 74 | 74 | 100 | 26 | 100 | 48 | 100 |
| 41-50 | 87 | 83 | 95.4 | 31 | 93.5 | 52 | 96.3 |
| 51-60 | 118 | 112 | 94.9 | 28 | 96.6 | 84 | 94.4 |
| 61-70 | 99 | 97 | 98.0 | 36 | 94.7 | 61 | 100 |
| 71-80 | 76 | 71 | 88.2 | 27 | 96.4 | 44 | 91.7 |
| 81-90 | 15 | 11 | 73.3 | 5 | 83.3 | 6 | 66.7 |
| Total | 564 | 538 | 95.4 | 189 | 96.3 | 373 | 94.9 |

For a further 46 patients, a total of 293 end-expiratory breath samples were analysed. The overall compliance rate is 88.1%, with a breakdown between male and female of 96.6% and 82.4% respectively. This compliance rate is comparable to the larger data set. The percentage of individual breath samples which achieved _*EE*_*O*_*2*_ below 14%, arranged by gender and age group is shown in Figure 3. When comparing the compliance rate with their Body Mass Index (BMI), samples delivered by patients with a BMI between 15 and 10 reduced to 72.6%, while samples from other BMI groups are all above 89%, Figure 4. There was no patient in the 36-40 BMI group in this study.

**Figure 3:**
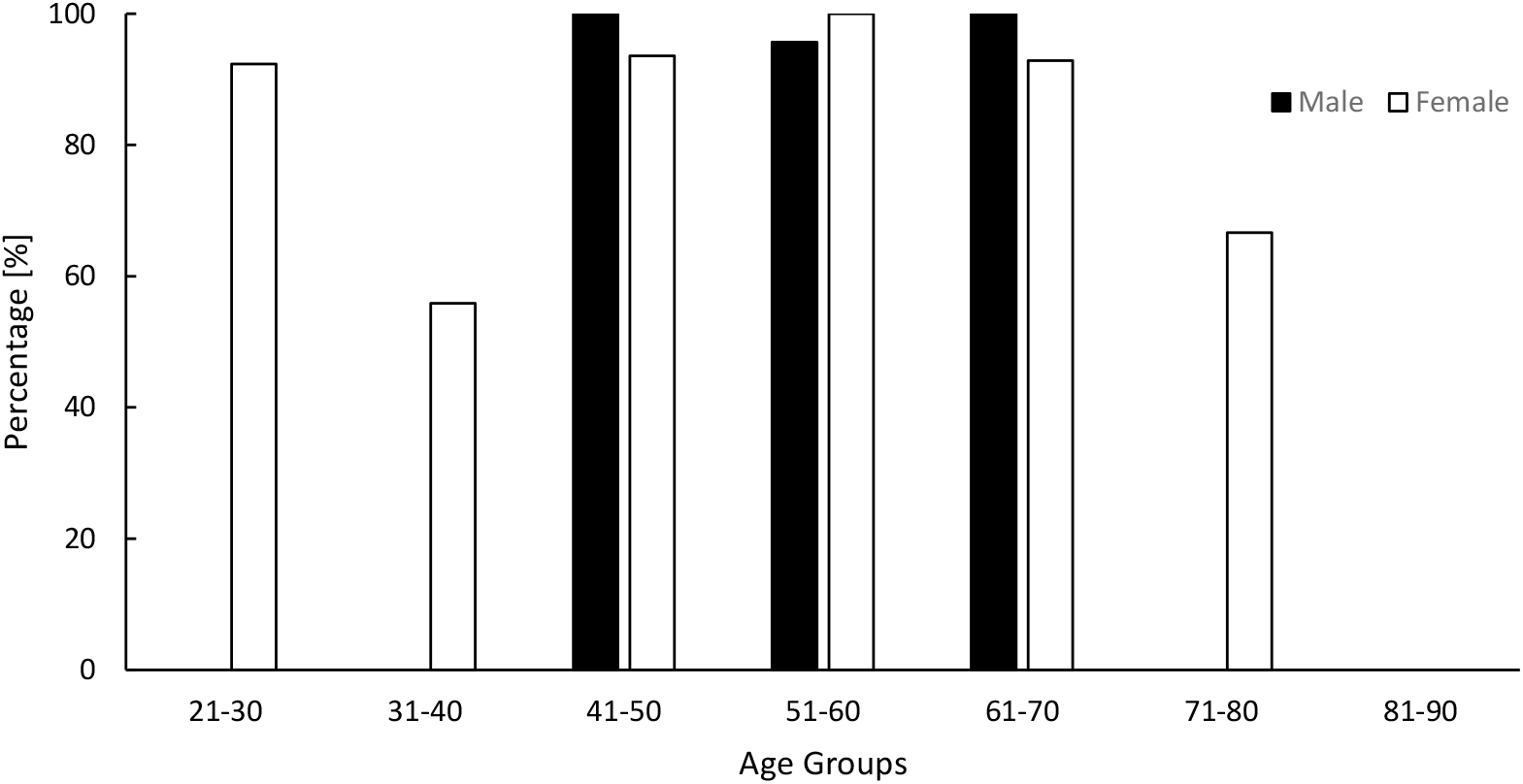
The percentage of individual breath samples which achieved _*EE*_*O*_*2*_ below 14% by gender and age group

**Figure 4:**
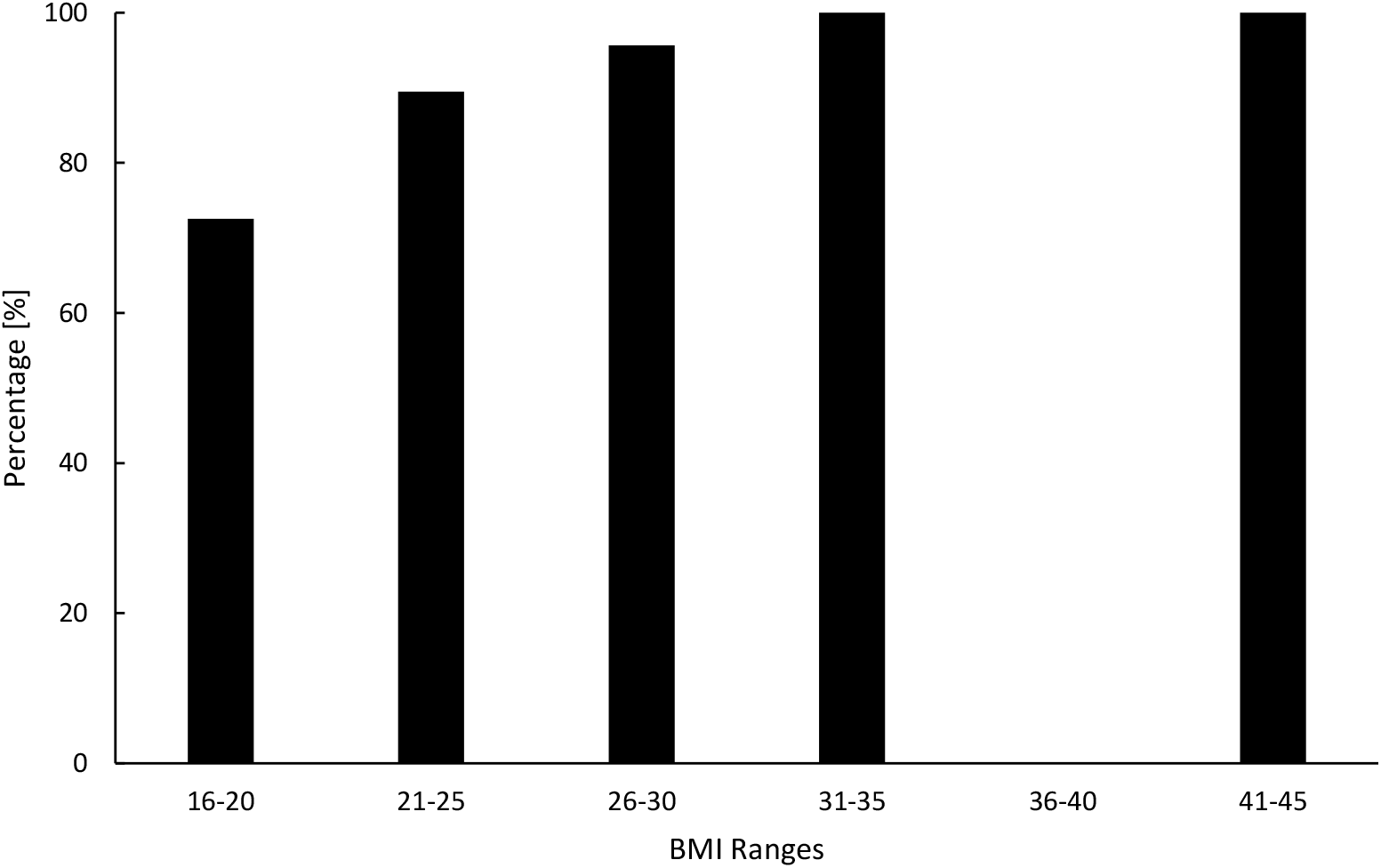
The percentage of individual breath samples which achieved _*EE*_*O*_*2*_ below 14% by patient’s BMI. (*Note: there was no patient in the data group of BMI 36-40 in this study*)

The results showed a wide range of _*EE*_*O*_*2*_, from the best sample of 9.5% to 16.2%. The mean _*EE*_*O*_*2*_ for the 293 samples is 12.9%, with a standard deviation of 1.1. The _*EE*_*O*_*2*_ data set also indicates a normal distribution, as shown in Figure 5.

**Figure 5:**
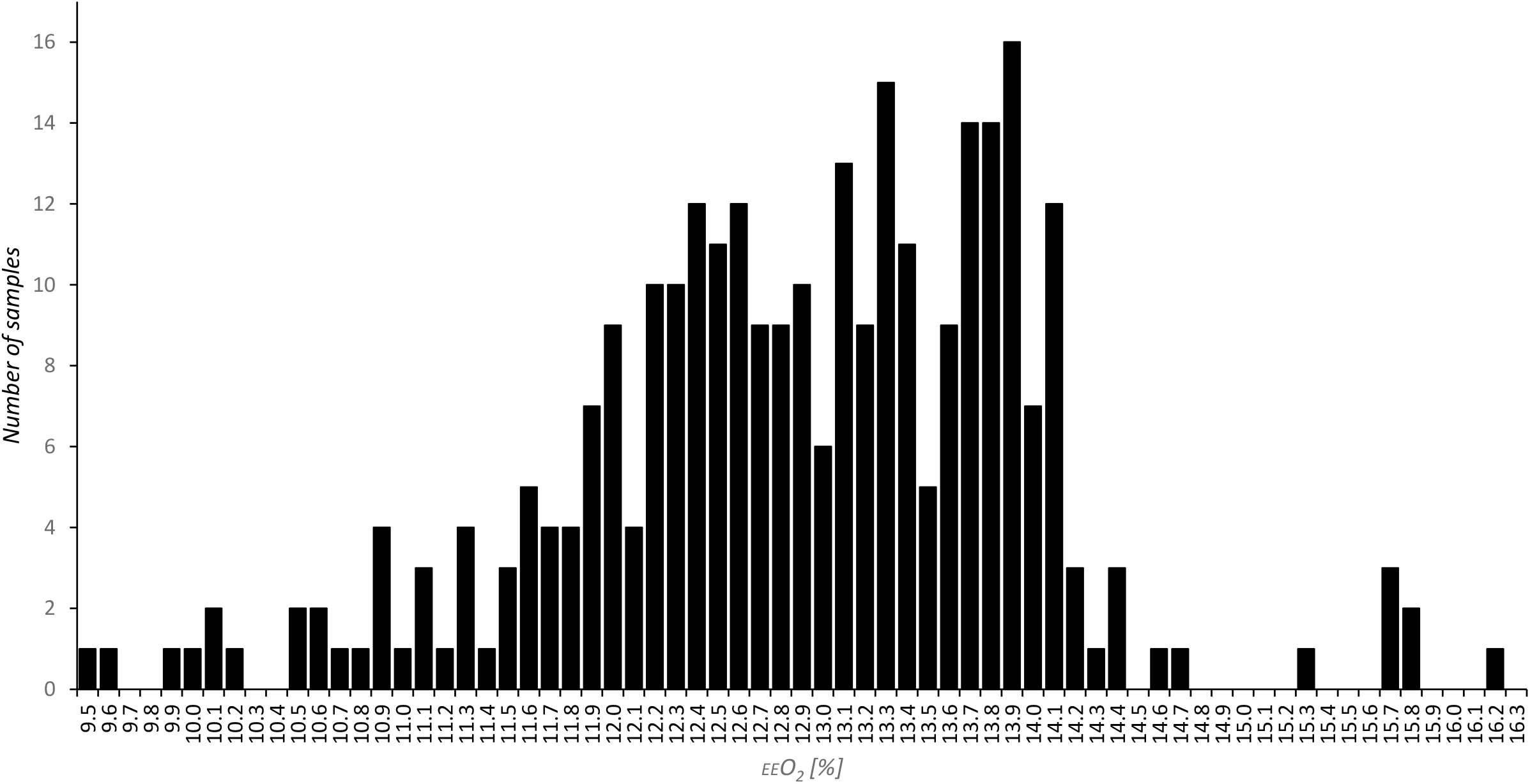
The distribution of _*EE*_*O*_*2*_ in end-expiratory breath samples

In order to evaluate the efficacy of the correction factor (CF), the end-expiratory hydrogen (_*EE*_*H*_*2*_) and end-expiratory methane (_*EE*_*CH*_*4*_) values were compared with the predicted hydrogen (_*CF*_*H*_*2*_) and predicted methane (_*CF*_*CH*_*4*_) values. The _*CF*_*H*_*2*_ and _*CF*_*CH*_*4*_ were calculated from the real-time H_2_and CH_4_ measurement when the O_2_concentration of the breath sample dropped to 15%. CF at 15% oxygen is 1.19. The range of difference recorded was between −30 ppm and 114 ppm.

The difference in percentage between the actual and the predicted values for H_2_ and CH_4_are shown in Figure 6a and 6b respectively. The results show the predicted values (_*CF*_*H*_*2*_ and _*CF*_*CH*_*4*_) often underestimated the actual measurements (_*EE*_*H*_*2*_ and _*EE*_*CH*_*4*_).

**Figure 6a:**
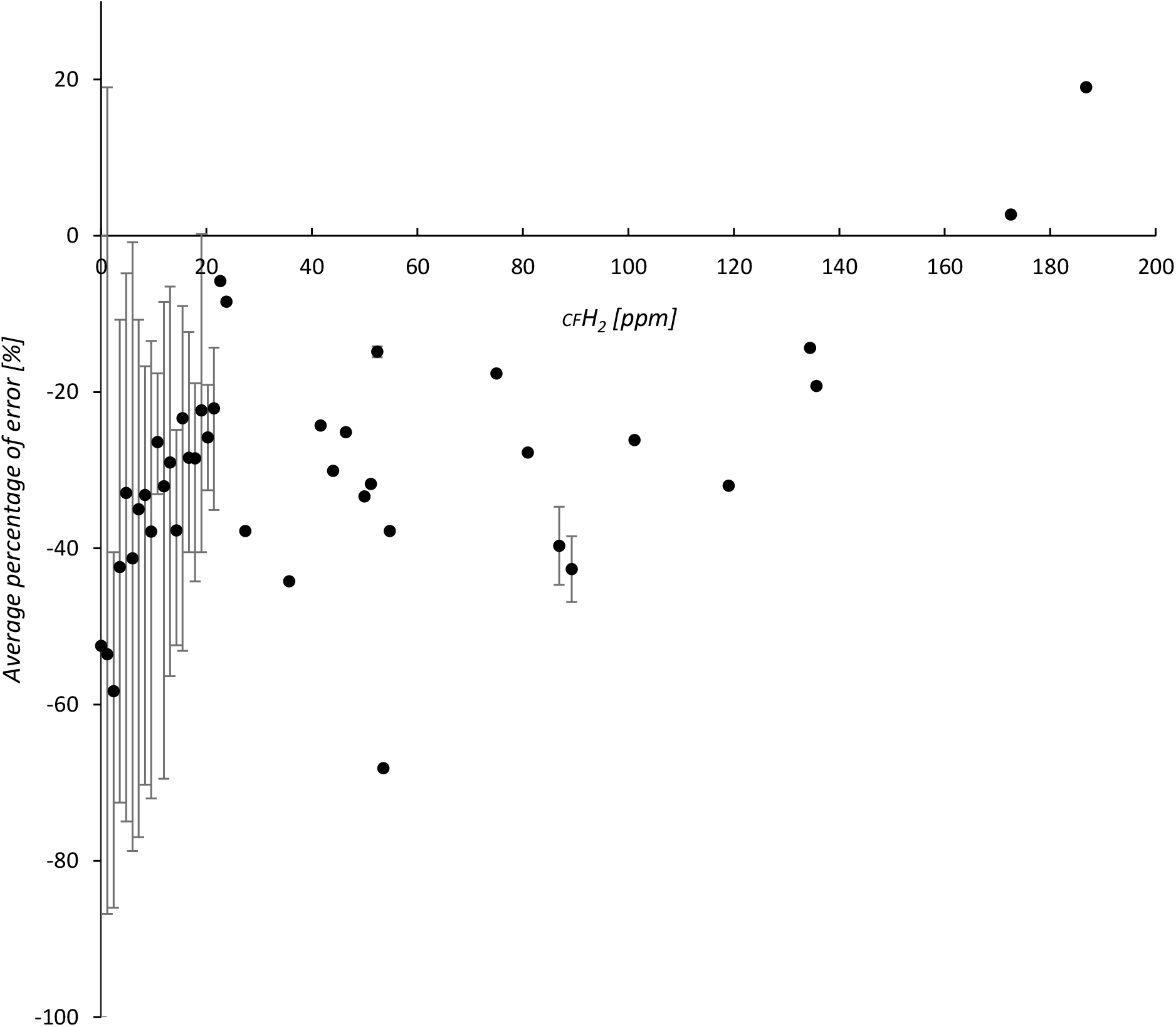
Percentage of error for _*CF*_*H*_*2*_ compared to _*EE*_*H*_*2*_.

**Figure 6b:**
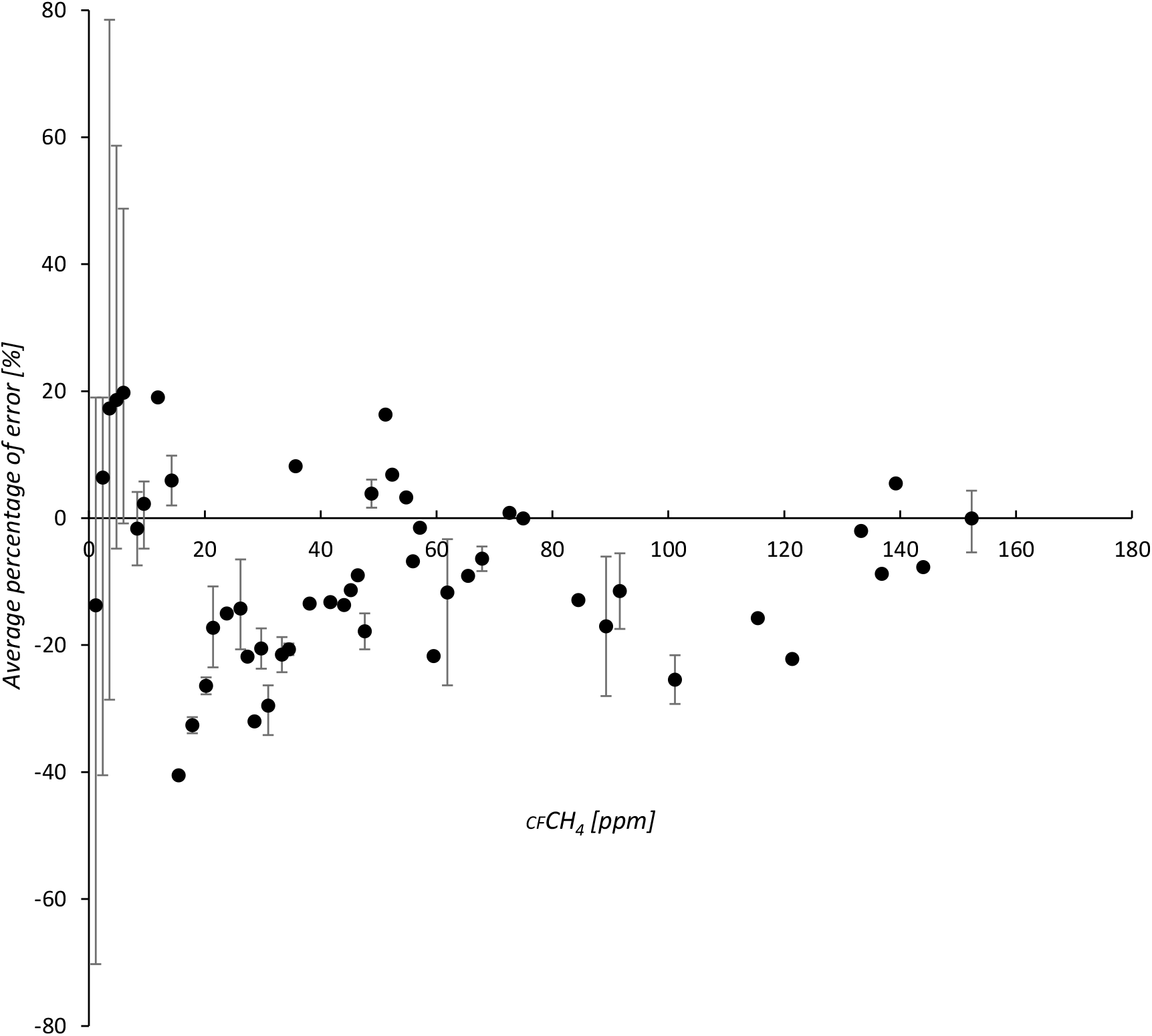
Percentage of error for _*CF*_*CH*_*4*_ compared to _*EE*_*CH*_*4*_.

The overall average difference for hydrogen measurement using the correction factor was −42.7%, while the average percentage error for methane measurement using the correction factor was 7.8%. However, it was noted that the percentage of error might be skewed when the measurement was at low level, e.g. _*CF*_*H*_*2*_ = 0 and _*EE*_*H*_*2*_ =1 will lead to 100% error. As the minimum threshold for positive result in GI Breath Test is 10ppm, the average error was adjusted by excluding any _*EE*_*H*_*2*_ below 10ppm. The adjusted average errors for _*CF*_*H*_*2*_ and _*CF*_*CH*_*4*_ were −36.4% and −12.8% respectively.

In spite of the large discrepancy between the end-expiratory measurement and the predicted value, majority of the cases arrived at the same diagnosis regardless of using either _*EE*_*H*_*2*_/_*EE*_*CH*_*4*_ or _*CF*_*H*_*2*_/_*CF*_*CH*_*4*_. There was no false positive CBT result found in the 46 cases in this study.

However, the analysis has revealed that the predicted values (_*CF*_*H*_*2*_or _*CF*_*CH*_*4*_) in 4 out of 8 positive cases led to a false negative result. This indicates a diagnostic error of 50%. Two such cases are shown in Figure 7a (Hydrogen false negative) and Figure 7b (Methane false negative) for a CBT (Glucose) test. They began with a high but stable baseline reading. Once glucose was administered, the _*EE*_*H*_*2*_ peaked within 20 minutes above the positive threshold. Both _*EE*_*H*_*2*_ and _*EE*_*CH*_*4*_ profiles indicated a typical glucose positive response. On the other hand, the predictive values (_*CF*_*H*_*2*_ and _*CF*_*CH*_*4*_) were unable to correctly predict the end-expiratory measurement and failed to indicate a positive result.

**Figure 7a:**
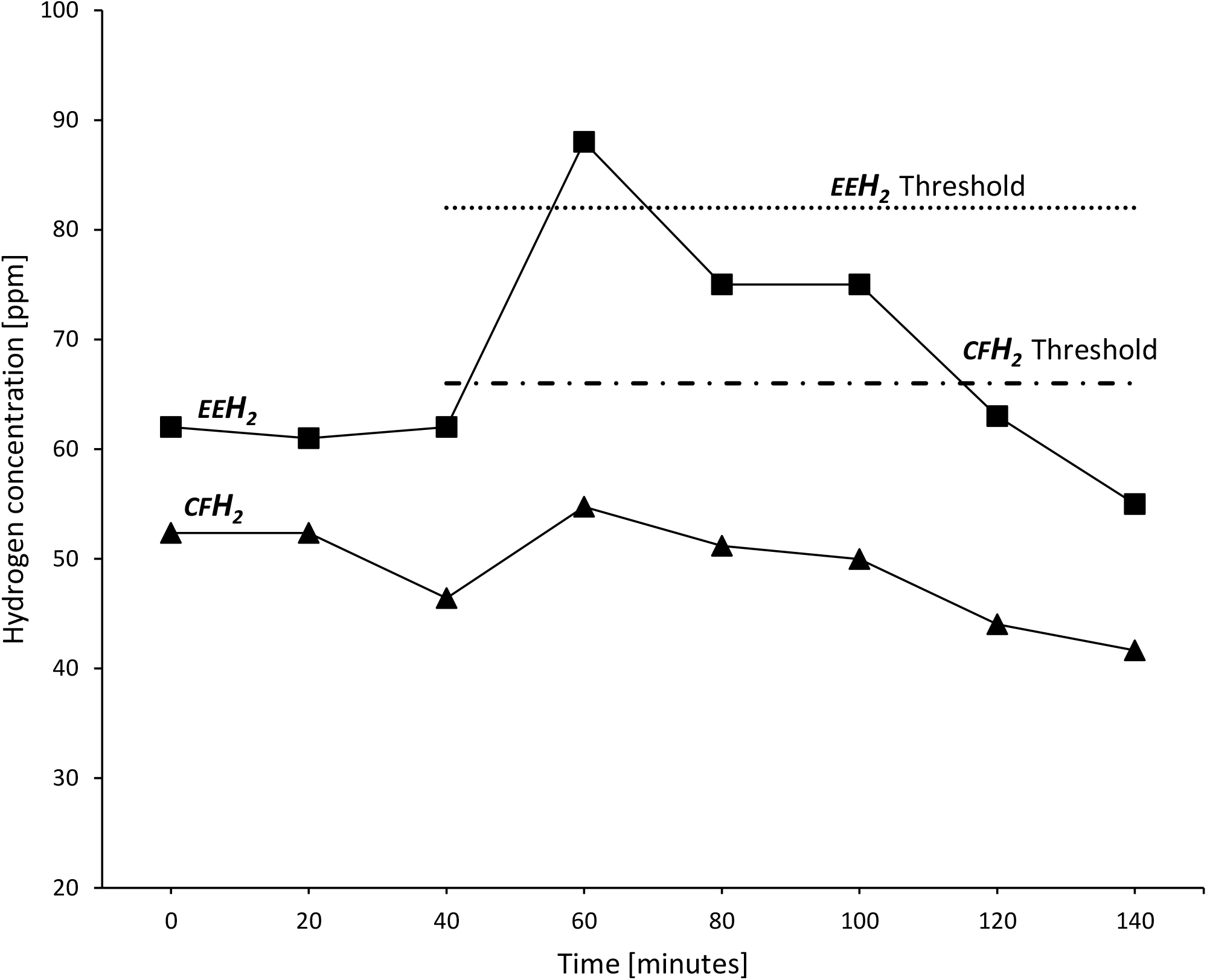
Difference between _*EE*_*H*_*2*_ and _*CF*_*H*_*2*_ values in a GI Breath Test (Glucose)

**Figure 7b:**
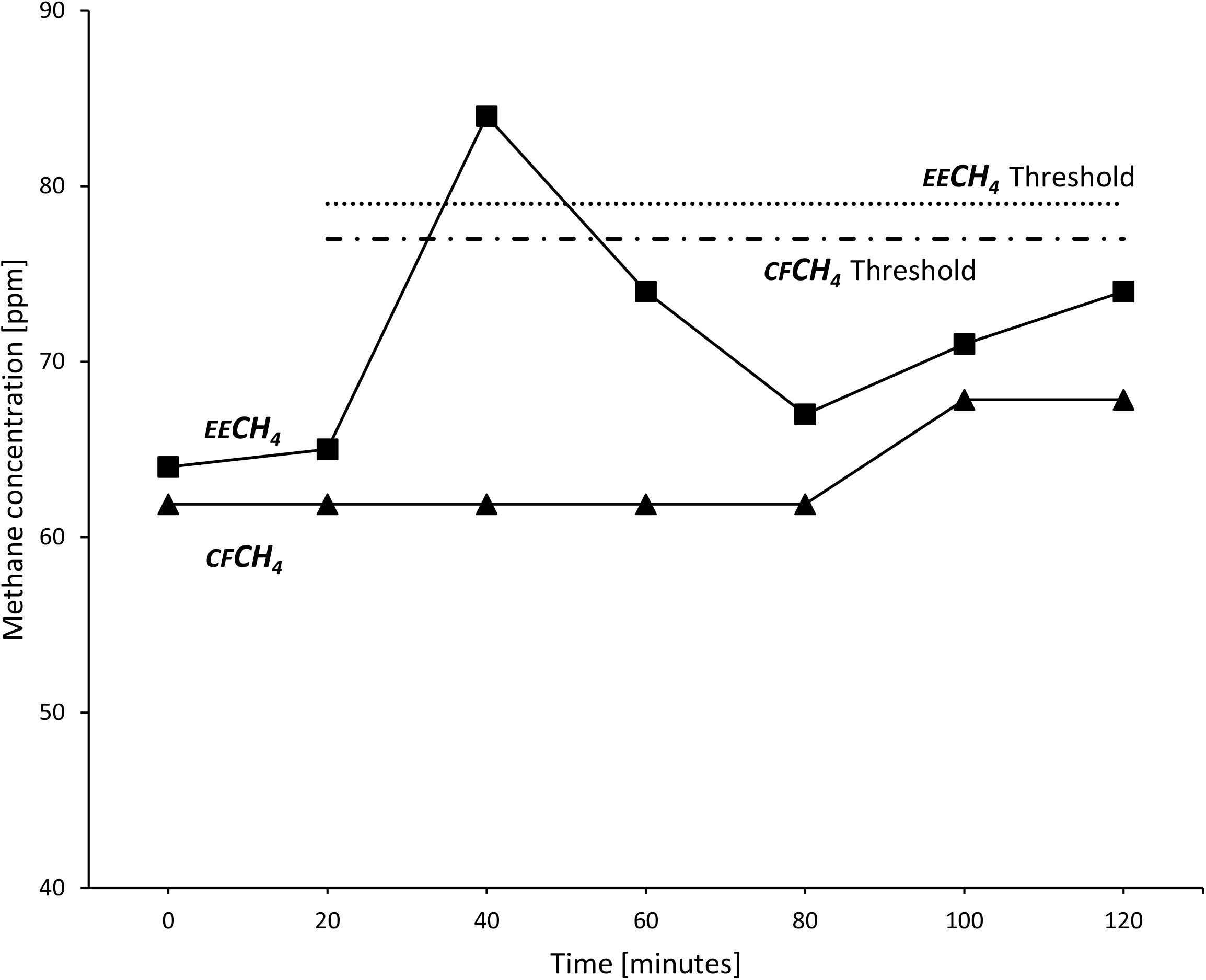
Difference between _*EE*_*CH*_*4*_ and _*CF*_*CH*_*4*_ values in a GI Breath Test (Glucose)

## DISCUSSION

The diagnostic result of the CBT depends greatly on the quality of the breath sample collected. Although it is not the only factor, it must not be underestimated. For CBT with real-time measurement, quality of the end-expiratory breath is critical to the accuracy of the hydrogen and methane level.

The results indicated that the majority of patients who have undergone CBT with real-time measurement are able to deliver an end-expiratory breath sample. End-expiratory Oxygen (_*EE*_*O*_*2*_) measured in this study indicated that it was usually better than the recommended 14%. The ability to deliver an end-expiratory breath was not affected by age, up to 80 years old, although there is an indication that the age group over 80 years old may have some difficulties. The compliance level did not appear to be affected by the patients’ gender. In comparison, BMI can have a more profound effect on the patient’s ability to deliver an end-expiratory breath sample.

Therefore, there is no obvious limitation or concern over the patient’s ability to provide an end-expiratory breath sample. It is imperative special care is taken to collect the breath samples and to ensure that patient breathes out completely into the analyser.

The additional 46 patients in the second part of the study were randomly selected in a continuous period of 12 weeks. The proportion of positive cases in this period is 17.4%. It is similar to the number of average positive cases over three years at the same centre, using the same breath analyser [16].

It was obvious from the analysis that _*EE*_*O*_*2*_ of the breath samples were not constant. In fact, it varied by a large degree, ranging from 16.2% to 9.5%. Nevertheless, the manufacturer has pre-defined the ideal or target _*EE*_*O*_*2*_ at 14%. The CF algorithm built into the GastroCH_4_ECK is similar to the method used in the carbon dioxide (CO_2_) correction factor employed in gas chromatography breath analysers [12]. The published data indicated the end-tidal breath should ideally contain 5% CO_2_ and suggested a specific correction factor algorithm which compensates for a range of CO_2_ levels between 2% −7%. The CF from the GastroCH_4_ECK only compensates oxygen level when the breath sample has an _*EE*_*O*_*2*_ level above 14%.

This study indicated that setting _*EE*_*O*_*2*_ at 14% is likely to be too high, as the mean _*EE*_*O*_*2*_ in this study was 12.9%. The rate of change in H_2_/ CH_4_values during an episode of breath sample collection will also significantly affect the validity of the CF and it expectedly varied to a large degree. Therefore, due to the highly inconsistent _*EE*_*O*_*2*_ and the varying nature of the H_2_/ CH_4_ in an episode of breath sample collection, the resulting _*CF*_*H*_*2*_/_*CF*_*CH*_*4*_ value based on _*EE*_*O*_*2*_ above 14% is often unreliable. Therefore, it is indeed possible to produce a false negative result if _*CF*_*H*_*2*_/_*CF*_*CH*_*4*_ is used.

As shown in this study, CF can contribute a level of uncertainty. When there is an operator who can ensure the quality of the breath sample during a real-time breath sample collection episode, CF may be redundant in real-time GI breath measurement.

On the other hand, O_2_ measurement is essential as a quality indicator for the breath sample. The continuous real-time trace of oxygen concentration on the GastroCH_4_ECK Gastrolyzer (Version 1) prevents the patient from accidentally breathing in during an episode of breath sample collection. Should secondary breathing be detected, the collection process can be aborted and re-started. This oxygen sensing feature ensures the quality of the breath sample by significantly reducing the uncertainties that arise from the sample collection stage and further exploiting the benefits of real-time breath measurement.

It may be important to note this study has focused on real-time breath measurement. Characteristics of O_2_ with a bag collection system are likely to be different and may require further study. In addition, the breath holding time has a significant effect to the mixture of gases in the end-expiratory breath [17]. Hence, breath sampling procedure must be designed to avoid the confusion between genuine maximum exhalation and the sensation of dyspnoea due to excessive breath hold.

## CONCLUSION

The majority of patients in this study were able to deliver breath samples below 14% O_2_. Oxygen concentration of an end-expiratory breath sample is largely unpredictable. The CF algorithm build-in to GastroCH_4_ECK proved to be unreliable and led to a diagnostic error of 50% of the positive cases in this study. Hence, with such uncertainty for the predicted H_2_/CH_4_values, it is essential that the actual end-expiratory breath is collected and CF may be redundant in real-time CBT. On the contrary, the on-screen continuous oxygen trace is highly valuable to ensure the quality of the end-expiratory breath sample collection.

## Data Availability

All data used in this study is openly available to the public, under the Freedom of Information Act 2000 in the United Kingdom.
The source data can be accessed via the web page of the Royal United Hospitals Bath NHS Foundation Trust. The specific address is provided in the section: Data Availability Links.

https://www.ruh.nhs.uk/MPB/documents/Service_data_2020b.pdf

## Notes

### Competing Interest Statement

The authors have declared no competing interest.

### Clinical Trial

The clinical data is publicly available.

### Funding Statement

The authors received no external funding for this work.

### Author Declarations

All data used in this study is openly available to the public, under the Freedom of Information Act 2000 in the United Kingdom.

### Summary of Updates

The number of individual samples studied increased from 228 to 293. The increased number of samples is now similar to the proportion of positive cases to a 3-year average at the same centre. Figure 3, 4 and 5 are updated to reflect the increased samples. Figure 6a, 6b, 7a and 7b are removed as it showed repeated information. The percentage of cases where it would have been recognised as false negative cases when Correction Factor is applied is 50%. A new graph added (Figure 7b) showing a typical false negative case from methane.

## REFERENCES

[1] Saad R J, Chey W D; Breath Tests for gastrointestinal disease: the real deal or just a lot of hot air?; Gastroenterology; 2007 Dec; 133(6): 1763–6. doi: 10.1053/j.gastro.2007.10.059

[2] Simrén M and Stotzer P-O; Use and abuse of hydrogen breath tests; Gut 2006 Mar; 55(3): 297–303. doi: 10.1136/gut.2005.075127

[3] Rezaie A, Buresi M, Lembo A, Lin H, McCallum R, Rao S, Schmulson M, VAldovinos M, Zakko S, Pimentel M; Hydrogen and Methane-Based Breath Testing in Gastrointestinal Disorders: The North American Consensus; Am J Gastroenterol; 2017; 112:775–784; doi: 10.1038/ajg.2017.46

[4] Gasbarrini A et al; Methodology and indications of H2-Breath testing in gastrointestinal diseases: the Rome Consensus Conference; Aliment Pharmacol Ther; 2009; 29 Suppl 1:1–49. doi: 10.1111/j.1365-2036.2009.03951.x

[5] Ghoshal U C; How to Interpret Hydrogen Breath Tests; J Neurogastroenterol Motil.2011 Jul; 17(3): 312–317. doi: 10.5056/jnm.2011.17.3.312

[6] Robb T A, Davidson G P; Advances in breath hydrogen quantitation in paediatrics: Sample collection and normalisation to constant oxygen and nitrogen levels; Clin. Chim. Acta; 1981; 111:281–5

[7] Hamilton L H;Breath Tests & Gastroenterology, Second Edition 1998.

[8] Lee W S, Davidson G P, Moore D J, Butler R N; Analysis of the breath hydrogen test for carbohydrate malabsorption: Validation of a pocket-sized breath test analyser; J. Paediatr. Child Health; 2000; 36:340–342

[9] Anderson J, Hlastala M P; Breath tests and airway gas exchange; Pulm Pharmacol Ther.2007; 20(2):112–7 doi: 10.1016/j.pupt.2005.12.002

[10] Fleming S C; Evaluation of a hand-held hydrogen monitor in the diagnosis of intestinal lactase deficiency; Ann Clin Biochem; 1990: 27:499–500

[11] de Lacy Costello B P J, Ledochowski M, Ratcliffe N M; The importance of methane breath testing: a review; J. Breath Res. 7; 2013 024001; doi: 10.1088/1752-7155/7/2/024001

[12] Niu H C, Schoeller D and Klein P. Improved gas chromatographic quantitation of breath hydrogen by normalization to respiratory carbon dioxide. J Lab Clin Med 1979; 94:755–63.

[13] Wolfson M R; Shaffer T H; Cardiopulmonary Physical Therapy (Fourth Edition), 2004–O2 14%

[14] Wagner P D; The physiological basis of pulmonary gas exchange: implications for clinical interpretation of arterial blood gases; European Respiratory Journal; 2015; 45:227–243

[15] GastroCH4ECK user manual (2014) Bedfont Scientific Ltd. UK

[16] Materacki L, Lee SM, Laidler P, Yong K, Betteridge F, Murugiah D, Colleypriest B; Is methane testing a useful adjunct to hydrogen breath testing?; BSG Annual Meeting 2018, Liverpool, UK, 4th – 7th June, 2018

[17] Levitt M D, Ellis C, Furne J; Influence of method of alveolar air collection on results of breath tests; Dig. Dis. Sci. 1998 Sep; 43(9):1938–45. doi:10.1023/a:1018874223418

